# Mortality during the COVID-19 pandemic: findings from the CLINIMEX exercise cohort in the year of 2020

**DOI:** 10.1101/2021.03.17.21253138

**Authors:** Claudio Gil S. Araújo, Christina Grüne de Souza e Silva, Claudia Lucia Barros de Castro, Jari A. Laukkanen, Jonathan Myers, Josef Niebauer, Aline Sardinha, João Felipe Franca

**Author notes:** Contact primary author: Dr. Claudio Gil S. Araújo.

## Abstract

**Background and Objective:** The COVID-19 pandemic has heavily hit Brazil and, in particular, our Clinic’s current location in Copacabana – Rio de Janeiro city, where, as of mid-February 2021, it led to one death per 266 inhabitants. After having recently updated the vital status and mortality data in our exercise population (CLINIMEX exercise cohort), we hypothesized that the review of their evaluation reports would offer a unique opportunity to unearth some relevant information about the association between selected variables assessed in our comprehensive Exercise Medicine evaluation protocol, in particular, aerobic and musculoskeletal (MUSK) fitness, clinical variables, and death due to COVID-19.

**Methods:** We conducted a retrospective study using data from the CLINIMEX exercise cohort that included 6,101 non-athletic men and women aged >30 years who were alive as of March 12^th^, 2020, and who’s vital status was followed up to December 14^th^, 2020. For data analysis, two approaches were used: 1) comparison of frequency of deaths and relative % of underlying causes of death between the last 18-months pre-pandemic and 9-month pandemic periods; and 2) data from 51 variables from the participant’s most recent evaluation, including sex, age and clinical profile plus other variables obtained from physical examination, spirometry, (MUSK) fitness (e.g., sitting-rising test) and maximal cycling leg cardiopulmonary exercise testing (e.g. maximal VO2 and cardiorespiratory optimal point) were selected for comparison between groups of non-COVID-19 and COVID-19 deaths. Results: Age at death varied from 51 to 102 years [mean = 80 years]. Only 4 participants that died – 3 COVID-19 and 1 non-COVID-19 - were healthy at the time of their evaluation [p=.52]. COVID-19 was the most frequent (n=35; 36.5%) cause of death among the 96 deaths during this 9-month period. Comparing pre-pandemic and pandemic periods, there was a 35% increase in deaths and proportionately fewer deaths due to neoplasia and other causes other than cardiovascular or endocrine diseases. Results of aerobic and MUSK fitness tests indicated that the majority of the study participants were relatively unfit when compared to available age and sex-reference values. Indeed, there were few differences in the 51 selected variables between the two groups, suggesting a somewhat healthier profile among COVID-19 death participants: lower body mass index [p=.04], higher % of predicted forced vital capacity [p=.04], lower number of previous percutaneous coronary interventions [p=.04] and lower resting supine diastolic blood pressure [p=.03], with no differences for aerobic/MUSK fitness variables or past history of exercise/sports [p>.05].

**Conclusion:** Our data support that COVID-19 was a frequent and premature cause of death in a convenience sample of primarily white, unhealthy, middle-age and elderly individuals and that data from exercise/sport history and physical fitness testing obtained some years earlier were unable to distinguish non-COVID-19 and COVID-19 deaths.

## Introduction

> ***We all know that death is inevitable but we do not know when it will occur and what its cause will be. Maybe knowing more about this might enable us to delay our death***

As of the current date (mid-February, 2021), more than 200 countries and territories have been affected by the COVID-19 pandemic, reaching extraordinary numbers of worldwide confirmed cases of >110 million affected individuals and >2.4 million deaths due to COVID-19, with 7-day moving averages slightly dropping from a recent peak of 750,000 new cases and 15,000 deaths per day.(1)

However, the impact of the COVID-19 pandemic, such as its temporal trends and its public health effects, varied throughout the different countries and regions around the world. For instance, whichever criteria is applied, the United States and Brazil emerged as the two most affected countries, both in absolute and relative (per capita) number of cases, hospitalizations and deaths. Moreover, even though they represent together only 6.9% of the world population, approximately one third of all confirmed cases and deaths from COVID-19 have occurred in either one of these two countries.(1) Therefore, the number of cases and deaths in both Brazil and the United States is four to five times higher than the world’s average, despite their widely recognized high-level quality of medical technology and human resources. (2-6)

Just over a year after the World Health Organization declared that COVID-19 was a public health emergency of international concern, a “coronavirus cascade” has occurred in scientific journals.(7) More than 100,000 COVID-19-related papers were published in PubMed and >10,000 in MedRvix. With the primary goal of sharing knowledge as fast as possible about the spread of the disease and its prevention and treatment, researchers around the world raced to share their work on COVID-19. However, it is particularly interesting that despite this large volume of papers, original scientific data about the relationship between COVID-19 and physical activity, exercise, sport practice and physical fitness are still scarce.(8-13) Since both cardiorespiratory or aerobic (14, 15) and musculoskeletal (MUSK) (16) fitness are positively and strongly associated with longer survival and several other favorable (but not all) health outcomes (17), it is plausible to speculate that individuals with higher fitness levels would have a more benign clinical course once infected by COVID-19. Indeed, there are very limited data on this matter and somewhat conflicting results (17-20). Moreover, to the best of our knowledge, there are no data regarding the potential association between exercise-related variables and the most relevant outcome, i.e., death from COVID-19.

This is of particularly interest as following strict recommendations from health authorities and institutions to stay home, maintain physical distancing and use mask, not surprisingly led to a significant drop in regular exercise and sports participation globally, contributing to a growing prevalence of sedentary lifestyle.(8, 9, 21-26) Even exercise-based cardiac rehabilitation programs were strongly affected, and thousands of exercise programs around the world had to interrupt their services either temporarily or permanently, or innovate with the help of modern technology and telemedicine resources in order to provide their services and keep their patients physically active.(27-30).

The first COVID-19 case in the city of Rio de Janeiro was recorded on March 6^th^, 2020, just 11 days after Brazil’s first registered case.(31) As of mid-February, COVID-19 mortality per one million inhabitants reached 1,810 in Rio de Janeiro state (17,366,189 inhabitants and 397 inhabitants/km^2^) increasing substantially to 2,700 in Rio de Janeiro city (6,747,815 inhabitants and 5,265 inhabitants/km^2^)(32) with a very high demographic density. However, as mentioned above, even in a single city such as Rio de Janeiro, huge inequalities can occur due to demographic characteristics, including population density and median age of inhabitants in given areas of the city.(31) For example, the 161,178 inhabitants of Copacabana (∼30% ≥60 years of age) have been extremely affected by the COVID-19 pandemic. In Copacabana, a famous upper middle-class and commercial coastline area, the inhabitants live primarily in apartment buildings that are literally packed in 7,84 km^2^ (including some green and mountain areas), with some statistics suggesting that the actual population density may reach the astonishing figure of 3.5 inhabitants/m^2^. Indeed, by again using mid-February/2021 for comparison purposes, Copacabana had registered 600 deaths, reflecting the tragedy of COVID-19 mortality of 3,750 per one million inhabitants, or 1 death per 266 inhabitants.

Located in one of the major streets of Copacabana (south zone of Rio de Janeiro city), the Exercise Medicine Clinic (CLINIMEX) is a privately-owned medical enterprise founded at the beginning of 1994, aiming to primarily provide medical services through comprehensive evaluation protocols and advanced medically-supervised exercise programs. Additionally, CLINIMEX conducts continuing education and has a strong interest and focus on developing innovations and high-quality research in the areas of exercise and health. As part of these actions, since its beginning, the CLINIMEX cohort study was planned and implemented, including all the participants who attended the Clinic for an Exercise Medicine evaluation and/or to attend a medically-supervised exercise program. Over these 27 years, the CLINIMEX cohort data with detailed information on aerobic or cardiorespiratory and non-aerobic or MUSK fitness have been carefully obtained and organized, allowing the research team – researchers and graduate students from different universities – to develop several scientific studies and to publish a number of papers and books at local, regional, national and international journals and/or publishers.

Periodically, as has recently occurred, vital status and mortality data of this cohort are updated. In reviewing a 9-month period, from mid-March/2020 – when Brazil had its first confirmed COVID-19 death – to mid-December/2020, a total of 97 deaths for different causes were identified in the participants of our cohort. Therefore, the analysis of the CLINIMEX cohort data provided a unique opportunity to analyze potential associations between the variables assessed in our comprehensive Exercise Medicine evaluation protocol, in particular, aerobic and MUSK physical fitness and clinical variables and death due to COVID19.

## Methods

### Study Sample

This is a retrospective study using the CLINIMEX open cohort that includes 9,243 participants evaluated at CLINIMEX from 1994 to December 14^th^, 2020. In brief, it is important to emphasize that this is a convenience and voluntary sample that substantially differs from the typical Rio de Janeiro’s city average population in terms of ethnic origin and socioeconomic status. While these items were not objectively assessed, it is assumed that almost all of the participants evaluated are in the upper quintile of the population for socioeconomic status and more than 90% are white. Moreover, despite the fact that about 10% are athletes(33) and nearly 20% of the participants were apparently healthy based on their baseline evaluation in the Clinic, the vast majority of the participants had chronic diseases, primarily cardiovascular diseases. Most of these participants were referred for evaluation at CLINIMEX by their own attending private physicians and the evaluations were paid by themselves and/or by their own personal/company medical insurance plan. Around 22% of the participants were evaluated more than once, comprising a total of 13,575 evaluations in these over 26 years. More details of the CLINIMEX cohort are provided in supplemental materials.

For the present study, a selected sample was obtained by applying the following exclusion criteria: 1) classified as an athlete,(33) 2) younger than 31 years of age at his/her first evaluation in the Clinic, and 3) deceased before Mar 12^th^, 2020. A detailed flowchart is presented in Figure 1. All participants read and signed an informed consent form authorizing the use of their de-identified data for research purposes. Retrospective mortality studies with the CLINIMEX cohort are officially registered and approved in the official Brazilian National Research Registry – Plataforma Brasil number CAEE 40122320.8.0000.9433 and were formally reviewed and approved by an external Research Ethics Committee – Centro de Capacitação Física do Exército (reference number 4.459.555, December 13, 2020) (https://plataformabrasil.saude.gov.br/login.jsf;jsessionid=8440F18B698A27430BBC0026F9C5C7F6.server-plataformabrasil-srvjpdf132).

**Figure 1.**
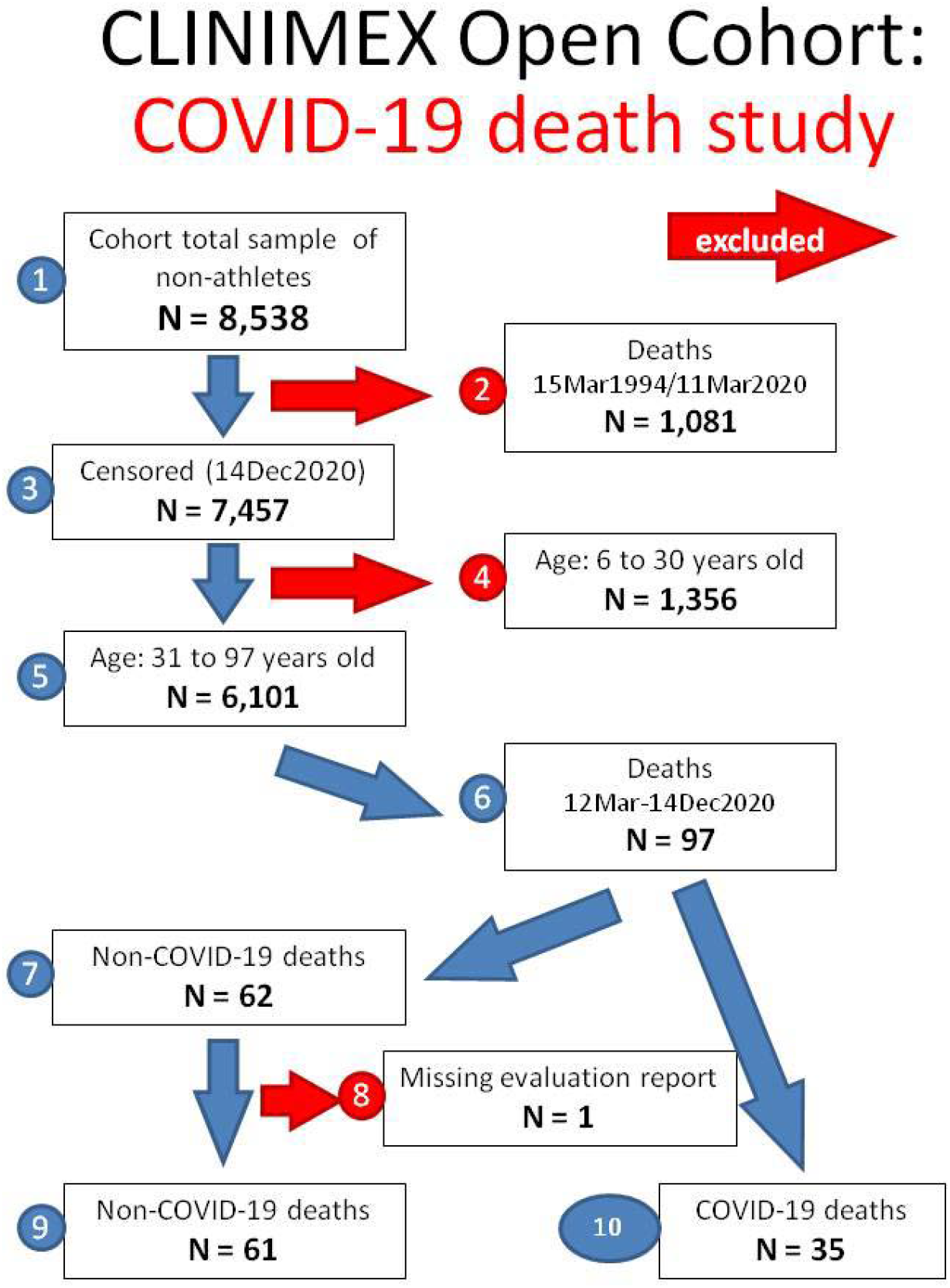
CLINIMEX open cohort: study flowchart. Blue and Red-numbered circles indicated each one of the steps to define the study sample. Red-numbered indicate exclusion points.

Censoring of vital status and mortality in the CLINIMEX cohort was updated to mid-December 2020 from the official registry data formally provided by the Rio de Janeiro State Secretary of Health. This information was complemented, when appropriate, by additional internet searches, CLINIMEX patient records and direct information obtained from assistant physicians and patients’ relatives and close friends.

After having applied the exclusion criteria, there remained a total of 6,101 alive participants – 4,004 men (65.6%) and 2,097 women (34.4%), aged 52.2 ± 12.7 years (mean ± standard deviation) at the time of their first evaluation. From these 6,101 participants, 97 (1.59%) died between March 12^th^ and December 14^th^, 2020, the first 9-month period of the COVID-19 pandemic in Rio de Janeiro, Brazil.

Using ICD-10 coding data, the main cause of death in 35 participants was COVID-19, and for the remaining 62 participants other non-COVID-19 causes. One of the non-COVID-19 deaths was excluded from the study due to a missing evaluation report file. Thus, the final study sample consisted of 96 patients who died during the first 9-month period of the COVID-19 pandemic in Brazil; these were divided into non-COVID-19 and COVID-19 deaths for comparison. Additionally, the CLINIMEX cohort death registry was searched for number and causes of death that occurred during the 18-month period preceding the pandemic (twice the period of time covered by this study), i.e., between mid-September, 2018 and mid-March, 2020, aiming to obtain information about the typical CLINIMEX cohort mortality pattern prior to the COVID-19 pandemic.

### CLINIMEX Evaluation Protocol

In brief, the CLINIMEX evaluation protocol starts with a clinical history – with special emphasis on physical activity and exercise/sport profile – followed by a physical examination. In sequence, several anthropometric measurements are collected and a standard 12-lead supine resting electrocardiogram is registered. As part of the evaluation protocol, participants undergo various assessment evaluations, including handgrip strength, maximal muscle power, Flexitest,(34) sitting-rising test and balance tests in order to assess musculoskeletal physical fitness - muscle strength/power, flexibility and balance-. Participants are then seated and properly fitted to a leg cycle ergometer (Inbramed CG-04 [Brazil] or Cateye EC-1600 [Japan]), and disposable electrodes are placed on their chest for monitoring and continuously recording a one-lead electrocardiogram (Micromed Wincardio [Brazil]). The evaluation continues with pulmonary function testing - lung spirometry – (Schiller [Switzerland] or Koko [United States] spirometers) and a cardiac vagal tone evaluation by the 4-second exercise test protocol. Finally, a maximal cycling cardiopulmonary exercise testing (CPX) is undertaken following an individualized ramp protocol based on the responses to the CLINIMEX Aerobic Questionnaire. The initial and increment in workload ratio are selected by the evaluator based on the information collected on the history of physical activity, exercise and sport in the most recent years. Participants expire through a pneumotach (MedGraphics prevent [United States]) adapted to a mouthpiece and measurements of air flow and expired gas fractions are carried out by a metabolic analyzer (MedGraphics VO2000 [United States]). The spirometer and metabolic analyzer are calibrated daily for flow and gas concentrations using a 3-L sealed syringe and standard gas mixtures. All tests are performed in an exercise lab at sea level with properly controlled temperature and relative humidity under the presence of qualified medical staff including at least one Exercise & Sports Medicine physician and a nurse assistant. Personnel were adequately trained and the exercise laboratory was equipped for handling medical emergencies that could occur during the evaluation protocol, as recommended by the Brazilian Society of Cardiology.(35)

### Selected Variables for the Study

From all variables and data collected during this comprehensive Exercise Medicine evaluation protocol, 51 variables were chosen for the comparison between the non-COVID-19 and COVID-19 death groups. A total of 12 variables collected either during the medical history or CPX were classified in binary format; sex (men or women), clinical status (healthy or unhealthy), and presence or absence (yes or no) of: coronary artery disease, myocardial infarction, previous percutaneous coronary intervention, previous coronary artery bypass grafting, arterial hypertension, dyslipidemia, diabetes mellitus, obesity, regular use of ß-blockers, and arrhythmias during maximal exercise testing.

The other 39 variables were divided in 7 groups:

- data derived from the censoring and from CLINIMEX registry: age at evaluation, age at death and elapsed time in days between evaluation and death;
- history of physical activity, exercise and sports practice in three different times of life: childhood and adolescence, adult life and prior year to evaluation (using an incremental ordinal scale from 0 to 4, where 0 refers to sedentary lifestyle, 1 represents insufficiently active [less than minimum recommended dose for age], 2 means achieving the age-recommended dose in terms of regular physical activity, exercise and/or sports, 3 reflects a very active lifestyle [exceeding the minimal recommended dose for age], and 4 is attributed for those that are exceeding the minimal recommended dose in several times and/or are involved in competitive sports that have a high aerobic requirement;(36)
- lung spirometry: forced vital capacity, forced expiratory volume in 1-second, peak expiratory flow and mid-expiratory flow (absolute and age and sex-predicted values);
- cardiac vagal tone evaluation: the ratio between two RR-interval durations measured in the electrocardiogram, the first one immediately before and the second one at the end of a 4-second fast and unloaded cycling test that is carried out in a full and maximal inspiratory apnea;(37, 38)
- anthropometry: height, weight, body mass index (a value ≥30 kg.m^-2^ used to classify participants as obese) and the sum of 4 skinfolds (triciptal, subscapular, suprailiac and abdominal);
- MUSK assessment: absolute and relative (to body weight) handgrip (absolute and relative to body weight),(39) maximal muscle power measured during upper row movement (absolute and relative to body weight),(40, 41) flexibility by Flexitest(34, 42) and the ability to sit and rise from the floor as assessed by the sitting-rising test;(16, 43)
- aerobic fitness assessment by maximal cycling CPX; heart rate and systolic and diastolic seated pre-exercise blood pressure values, maximal and 1-min post-exercise heart rate, maximal systolic and diastolic blood pressure, test duration, oxygen uptake at ventilatory anaerobic threshold (VO2 AT), maximum oxygen uptake (VO2 max) and cardiorespiratory optimal point – the minimal mean ventilation/oxygen uptake in a given minute of CPX -(44, 45) (absolute or relative to body weight and age and sex-predicted values depending on the variable assessed) (please access the supplemental material for a more detailed description of the selected variables for this study and (more detail on the CLINIMEX evaluation protocol).

### Statistical analysis

Descriptive statistics are presented as number of participants, mean, and standard deviations for continuous variables and as frequency and percentage for nominal variables. Statistical comparisons between the two groups were made by unpaired two-tailed t-test and chi-square test, respectively, for continuous and nominal variables. A 5% level of probability was set for statistical significance. Inferential testing results with probabilities between 5 and 10% were considered as borderline in terms of statistical significance. Prism software version 8.4.3 (GraphPad, San Diego, US) was used for all statistical calculations.

## Results

Considering the 9-month period of the study, 96 of the 6,101 participants died, corresponding to 1.6% of the study’s sample. COVID-19 was the most frequent underlying cause of death in our cohort representing 35 participants and 36.5% of all 96 identified deaths or 0.57% of the total selected cohort participants for this study. In order to place this information in context, 35 COVID-19 deaths represents a mortality ratio of 5,736/one million or 1 death/174 cohort participants, corresponding to >20 times and >1.5 times, the world’s average and Copacabana’s mortality ratio respectively, when compared here with data from the end of 2020. Among the other specific causes of death, cardiovascular diseases, neoplasia and respiratory diseases were, in sequence, the most common underlying causes of death.

Considering the typical cohort historical profile of deaths as ascertained from the pre-pandemic 18-month period, there were clear differences. First, the absolute number of deaths increased by 35% comparing the 9-month (obtained by dividing the 18-month total value by 2) pre-pandemic and pandemic periods (71 versus 96 deaths). While no deaths were previously reported from COVID-19, there were also differences in the profile of underlying causes of deaths between the two periods of time considered. Since the fraction of deaths due to cardiovascular and endocrine diseases were quite similar during the two studied periods –28.2% and 4.2% (pre-pandemic) versus 29.2% and 3.1% (pandemic) -, the percentage of participants dying from causes such as neoplasia, respiratory, digestive and renal diseases, were clearly higher (2-3x) during the pre-pandemic period. Table 1 presents detailed information about the underlying causes of death during the two periods of time analyzed.

**Table 1.** CLINIMEX cohort: comparing primary causes of deaths in two periods of time^*^

|  | Pre-pandemic period |  | Pandemic period |  |
| --- | --- | --- | --- | --- |
|  | N | (%) | N | (%) |
| TOTAL | 142 | 100.0 | 96 | 100.0 |
| 1 COVID-19 | 0 | 0.0 | 35 | 36.5 |
| 2 Cardiovascular diseases | 40 | 28.2 | 28 | 29.2 |
| 3 Neoplasia | 30 | 21.1 | 9 | 9.4 |
| 4 Respiratory diseases | 26 | 18.3 | 6 | 6.3 |
| 5 Endocrine diseases | 6 | 4.2 | 3 | 3.1 |
| 6 Digestive diseases | 9 | 6.3 | 2 | 2.1 |
| 7 Neurological diseases | 5 | 3.5 | 2 | 2.1 |
| 8 Renal diseases | 5 | 3.5 | 1 | 1.0 |
| 9 Sepsis | 9 | 6.3 | 3 | 3.1 |
| 10 Others | 3 | 2.1 | 2 | 2.1 |
| 11 External causes | 7 | 4.9 | 3 | 3.1 |
| 12 Undetermined | 2 | 1.4 | 2 | 2.1 |

\* according to ICD-10 in the official death certificates

The first cohort COVID-19 death occurred on April 3^rd^, 2020. The month-to-month analysis indicated that non-COVID-19 and COVID-19 deaths peaked in May/2020 during the 9-month period of study, with 10 cases for each group. Indeed, from April 23^rd^ to May 22^nd^, 2020(a 30-day period), 14 participants died from COVID-19 as compared to 5 participants dying from other causes. Two-thirds of COVID-19 deaths occurred in the months of April, May and June/2020, that is, in only three of the 9 months of study observation. By carefully looking at the non-COVID-19 and COVID-19 deaths on a monthly basis, it was possible to determine that, indeed, the frequency of monthly deaths dramatically changed over the 9-month period considered in the study (Figure 2).

**Figure 2.**
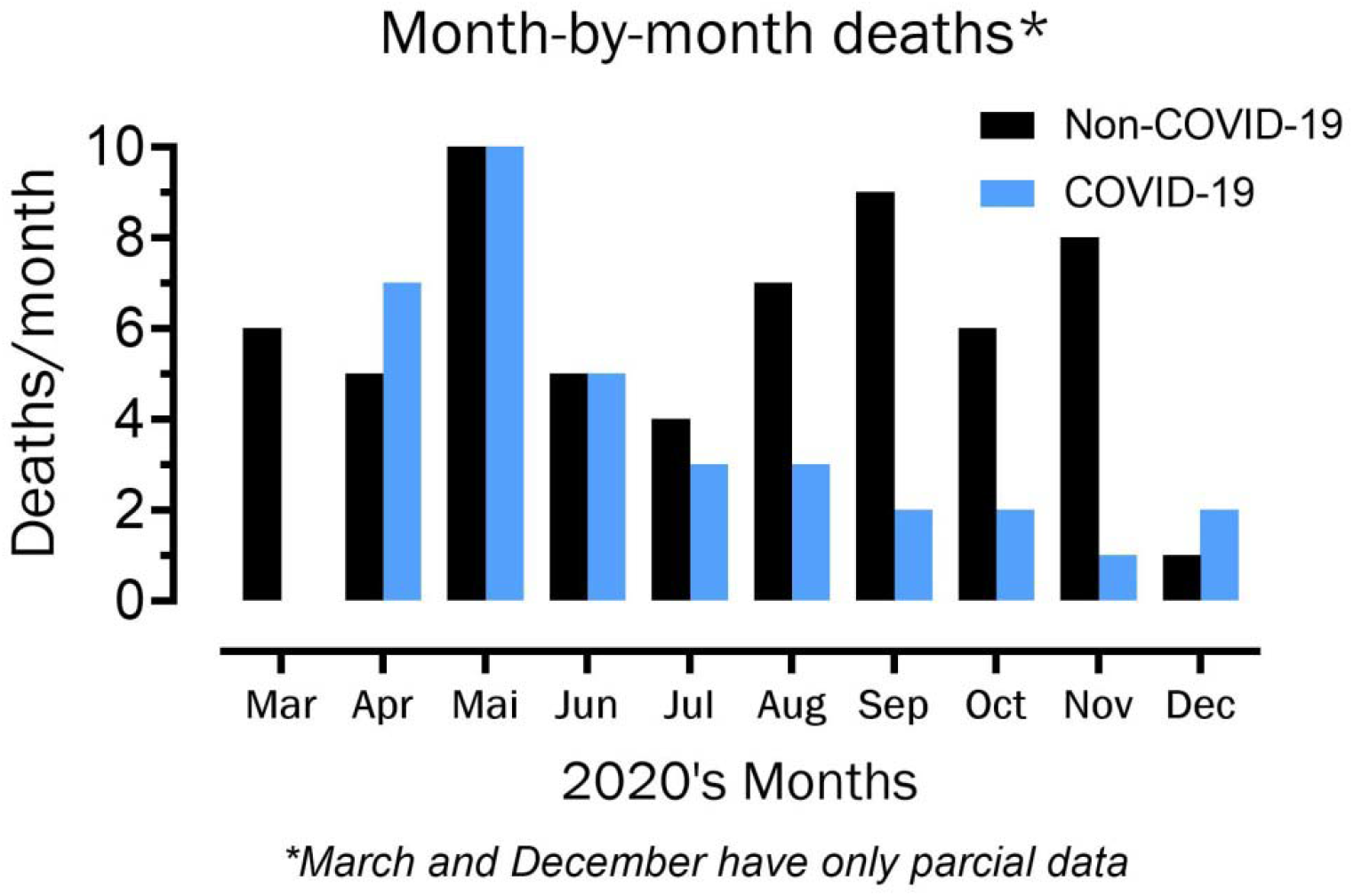
Month-by-month non-COVID-19 and COVID-19 deaths in the 9-month period of study observation

The main results of descriptive and inferential statistics for all 51 variables in both study groups are presented in tables 2 (binary variables) and 3 (continuous variables). Since there were missing data for some variables, the exact number of subjects in both groups for all variables is also shown. Sex distribution was similar between the groups of participants with COVID-19 and non-COVID-19 deaths with women comprising 22% and 20% in each group, respectively [p=.913]. This permitted us to combine participants of both sexes within each group for subsequent analyses. Regarding clinical status, almost all – 92 (96%) – of the participants that died during the 9-month period were classified as unhealthy according to the results of their last evaluation carried out some years before their death, with no clear distinction between the two groups [p=.269].

**Table 2.**
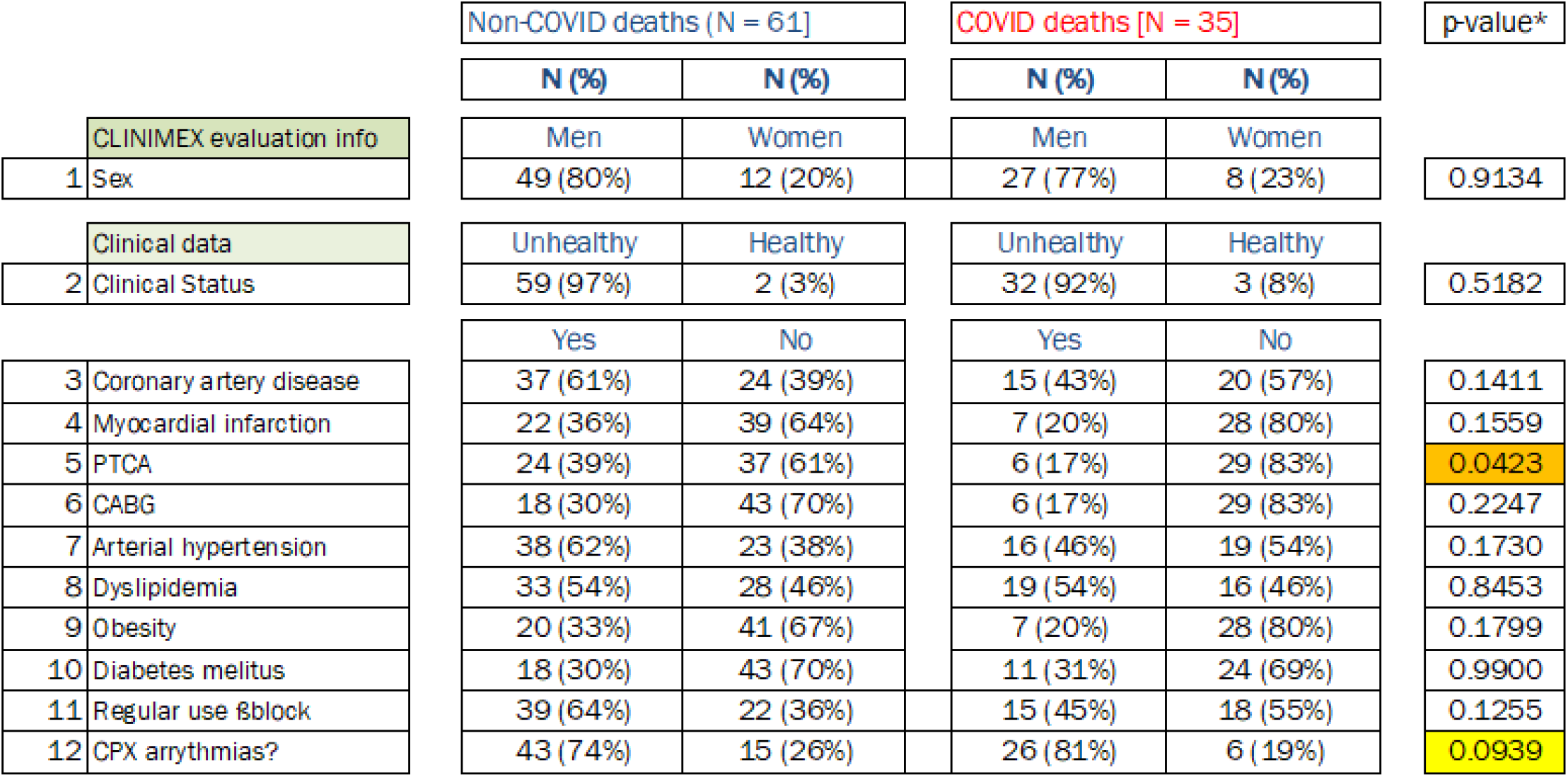
Main demographic and clinical data from the 96 participants from the CLINIMEX cohort who died between March 12th and December 14th, 2020, according to COVID-19 or non-COVID-19 cause of death

|  |  | Non-COVID deaths (N = 61] |  | COVID deaths [N = 35] |  | p-value* |
| --- | --- | --- | --- | --- | --- | --- |
|  |  | N (%) | N (%) | N (%) | N (%) |  |
| CLINIMEX evaluation info |  | Men | Women | Men | Women |  |
| 1 | Sex | 49 (80%) | 12 (20%) | 27 (77%) | 8 (23%) | 0.9134 |
| Clinical data |  | Unhealthy | Healthy | Unhealthy | Healthy |  |
| 2 | Clinical Status | 59 (97%) | 2 (3%) | 32 (92%) | 3 (8%) | 0.5182 |
|  |  | Yes | No | Yes | No |  |
| 3 | Coronary artery disease | 37 (61%) | 24 (39%) | 15 (43%) | 20 (57%) | 0.1411 |
| 4 | Myocardial infarction | 22 (36%) | 39 (64%) | 7 (20%) | 28 (80%) | 0.1559 |
| 5 | PTCA | 24 (39%) | 37 (61%) | 6 (17%) | 29 (83%) | 0.0423 |
| 6 | CABG | 18 (30%) | 43 (70%) | 6 (17%) | 29 (83%) | 0.2247 |
| 7 | Arterial hypertension | 38 (62%) | 23 (38%) | 16 (46%) | 19 (54%) | 0.1730 |
| 8 | Dyslipidemia | 33 (54%) | 28 (46%) | 19 (54%) | 16 (46%) | 0.8453 |
| 9 | Obesity | 20 (33%) | 41 (67%) | 7 (20%) | 28 (80%) | 0.1799 |
| 10 | Diabetes melitus | 18 (30%) | 43 (70%) | 11 (31%) | 24 (69%) | 0.9900 |
| 11 | Regular use βblock | 39 (64%) | 22 (36%) | 15 (45%) | 18 (55%) | 0.1255 |
| 12 | CPX arrhythmias? | 43 (74%) | 15 (26%) | 26 (81%) | 6 (19%) | 0.0939 |

In terms of the remaining clinical data, table 2 shows that participants who died from non-COVID-19 causes exhibited a worse profile for most of these variables with a significant difference observed for history of previous percutaneous coronary intervention [p=.0423]. Presence of coronary artery disease, arterial hypertension, obesity and regular use of ß-blockers, as well as history of myocardial infarction and coronary artery bypass grafting were 1.5 to 1.9 times more common among participants that died from non-COVID-19 causes, although these differences did not reach statistical significance [p values ranging from .1255 to .2247]. Nevertheless, nearly identical percentages were found for participants in both groups for presence of dyslipidemia and diabetes mellitus; 54% and 30% respectively. On the other hand, borderline statistical significance was observed for the presence of any type of arrhythmias during or immediately after CPX [p=.0939], with a slightly lower percentage among non-COVID-19 as compared to COVID-19 participants, respectively, 74% versus 81%.

The main findings - mean, standard deviations and number of cases -, as well as the corresponding statistical probability level found for the comparisons between the two groups for the 39 continuous variables studied are presented in table 3 and further described as follows. Age at last evaluation, age at death and time elapsed from evaluation to death was remarkably similar between non-COVID-19 and COVID-19 groups [p values ranging from .87 to .99]. Similarly, no differences were found for history of physical activity, exercise and sport in childhood/adolescence or adult life and the prior year of the evaluation [p values ranging from .43 to .78]. On the other hand, despite very similar heights – mean difference of only 1.2 cm -, the participants dying from non-COVID causes were 7 kg heavier [p=.0500], resulting in a 3 kg.m^2^ higher body mass index – 28.9 versus 26.9 [p=.04] and a 13% higher sum of the 4 skinfolds [p=.11] when compared to those who died from COVID-19.

**Table 3.**
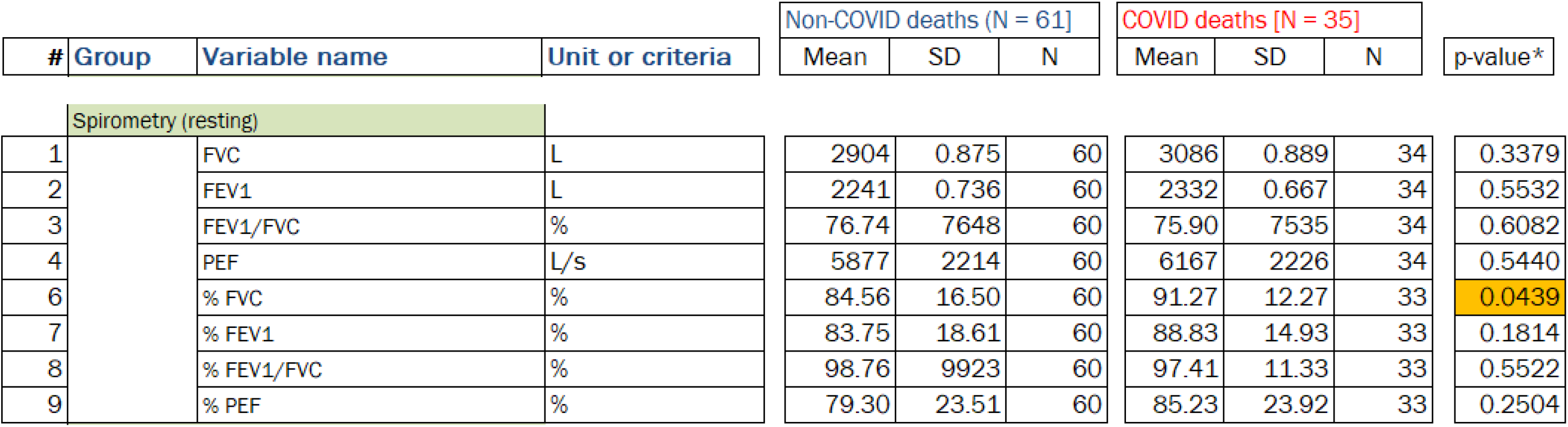
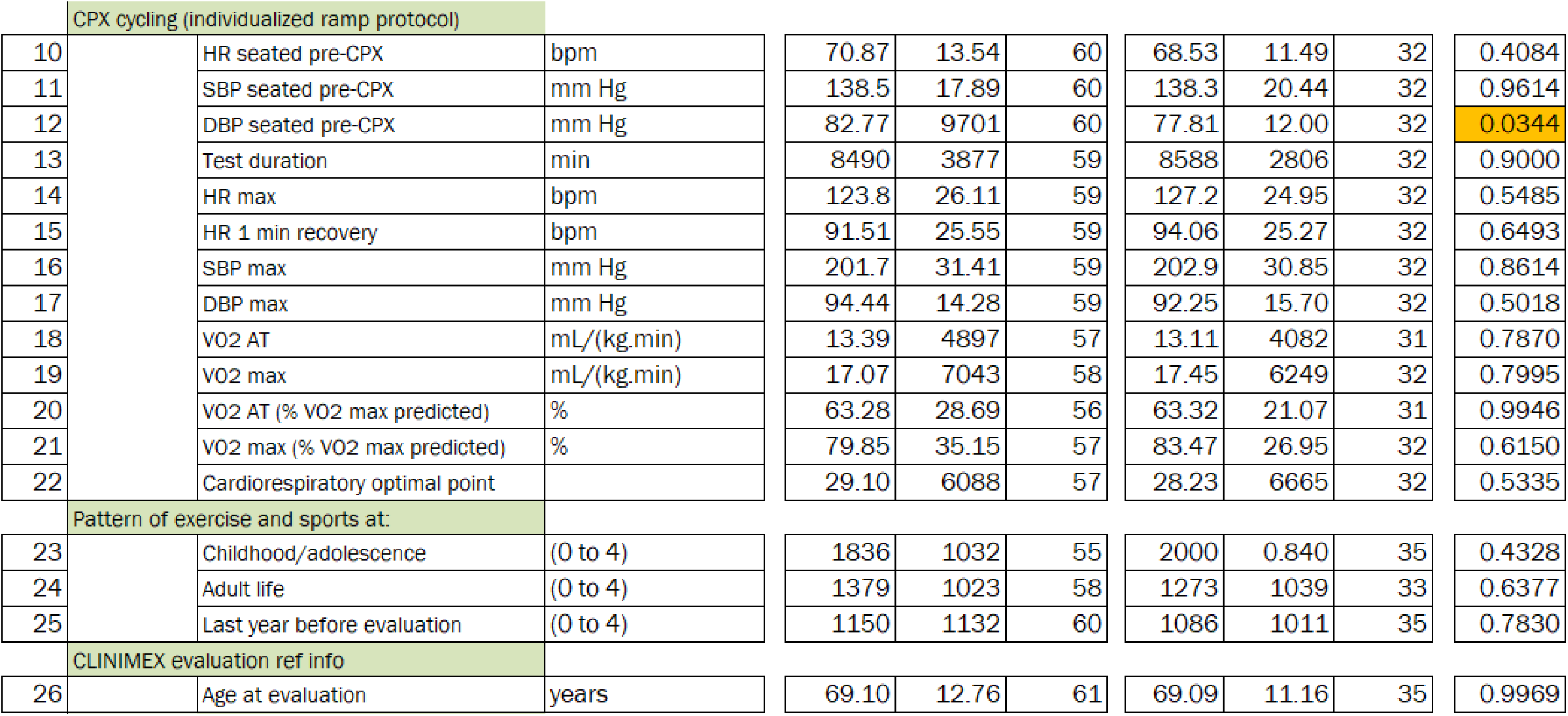
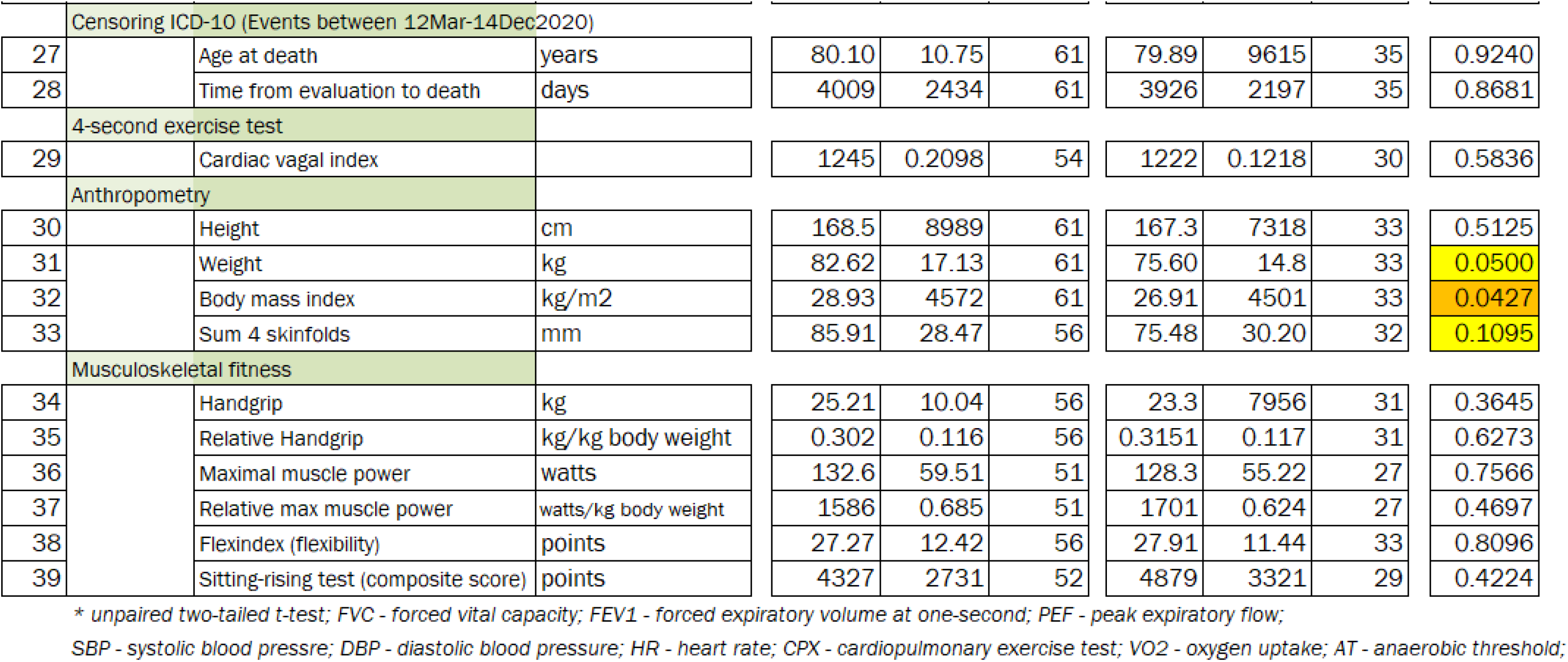
Results from the physical fitness assessment through the CLINIMEX evaluation of the 96 partici pants from the CLINIMEX cohort who died between March 12th and December 14th. 2020. according to C0VID-19 or non-C0VID-19 cause of death

Concerning resting lung spirometry, participants from the two groups showed mean results for 3 of 4 variables that were slightly lower than age- and sex-predicted, with a significantly lower value for % predicted forced vital capacity in non-COVID-19 when compared to the COVID-19 death group (85% versus 91%, respectively [p=.0439]). Results of the 4-second exercise test were similarly lower than expected, corresponding to the 38^th^ percentile when age-adjusted in the two groups [p=.58].

Data from 13 continuous variables derived from the CPX were analyzed involving cardiovascular and respiratory variables. Similar mean values were found for seated pre-CPX heart rate [p=.41] and systolic blood pressure [p=.96] in the two groups, with a 5-mm Hg lower mean seated pre-CPX diastolic blood pressure [p=.0344] for COVID-19 death participants. Overall median CPX duration was 9 minutes, with most of the participant’s CPX lasting between 8 and 12 minutes; CPX duration was almost identical between the two groups [p=.90]. Hemodynamic exercise data – maximal systolic and diastolic blood pressure, maximal heart rate and 1-min post-exercise heart rate were not different between those dying from non-COVID-19 and COVID-19 causes [p values ranging from .50 to .86].

In analyzing CPX ventilatory data, there were absolute and relative to body weight results. In general, the 96 participants that died showed poor aerobic fitness, with a mean VO2 max of only 17 mL.kg-1 .min-1 [<5 METs] that corresponds to a mean of only 63.3% of age- and sex-predicted VO2 max. There were no significant differences between non-COVID-19 and COVID-19 death groups for any of the five aerobic-related variables studied [p values ranging from .53 to .99].

Several test results were obtained from individual components or from overall MUSK fitness. There were no statistical differences between non-COVID-19 and COVID-19 death groups for any of the 6 MUSK variables assessed [p values ranging from .26 to .81]. Generally, overall mean MUSK results were clearly lower than expected based on age- and sex-predicted values – around the 20^th^ percentile with the exception of the Flexindex(34, 42) (the sum of the scores obtained from 20 different joint movements in a single participant) which was approximately the 30th percentile.

## Discussion

This study is, to the best of our knowledge, the first to address the relationship between physical fitness and exercise-related variables and COVID-19 deaths, by retrospectively reviewing and combining data from a well-established exercise-oriented epidemiological cohort and an official death certificate registry. We found that COVID-19 was the most frequent and premature cause of death in this convenience sample of primarily white and middle-aged and elderly participants with chronic non-communicable diseases in the CLINIMEX cohort (Rio de Janeiro – Brazil). Additionally, exercise/sport history and aerobic and MUSK physical fitness testing data obtained some years earlier were unable to distinguish between non-COVID-19 and COVID-19 deaths.

As currently documented in United States(2) (and perhaps in other countries as well), COVID-19 was the leading cause of death during the first 9-months of Brazil’s pandemic period in primarily chronically ill middle-aged or elderly men and women belonging to the CLINIMEX cohort that have been evaluated along the 26 years of existence of the Clinic.

Interestingly, the month-by-month analysis indicated that COVID-19 and non-COVID-19 deaths peaked in May/2020 (a total of 20), a number not reached previously in the entire cohort’s history during the 9-month period of observation. However, the death profile substantially changed over these 9 months. While most of the COVID-19 deaths occurred between April and June/2020, the non-COVID-19 deaths were more uniformly distributed along the full period of observation. An exception occurred in May/2020 when non-COVID deaths reached their peak; coincidently this was the peak of the first COVID-19 wave in the city of Rio de Janeiro and when the entire City’s public and private health systems were overstressed and had nearly collapsed. Among several explanations, two of them are highly likely: 1) Were some of these non-COVID-19 deaths in fact COVID-19-related deaths that were misclassified in the death certificate? 2) Was the overcrowded city’s health system unable to provide adequate services to patients who would not have typically died and they either died at home or from insufficient access to expected and needed medical services?

There are several possible reasons to explain why COVID-19 deaths were not evenly distributed over the 9-month period. Perhaps the most important is the fact that this profile somewhat reflects the total of COVID-19 deaths in Rio de Janeiro city and was likely influenced by a combination of high transmission rates, very limited testing, poor contact tracing strategies, poor adherence to physical distancing and mask use and potentially ineffective medical therapeutic choices (e.g., hydroxycloroquine and azythromicyn). All of these factors might have contributed to the proportionally higher mortality ratio seen during the initial months of the COVID-19 pandemic. Nevertheless, in contrast to most countries in the world, Rio de Janeiro city has not seen a period of strong pandemic deceleration of COVID-19 infection or hospitalizations, and deaths have continued to occur through this writing (mid-February/2021), as is partly reflected by our updated data. Additionally, while our data suggest that some underlying causes of death were proportionally less common during the pandemic, the small absolute number of deaths for each specific cause surely precluded a more definitive analysis of this information in our study. Future detailed epidemiological studies are recommended to address and clarify these relevant questions.

Our study benefits from the fact that sex distribution, age at evaluation, age at death and time elapsed between these two events were not significantly different between those dying from non-COVID-19 and COVID-19 causes. Only 3 of the total 96 deaths – 2 non-COVID-19 and 1 COVID-19 - were younger than 60 years old. Due to the rather restricted number of deaths, this study was not able to deeply analyze the demographic and clinical profiles of participants that died from COVID-19, in contrast to other much larger clinical and epidemiological studies.(46-52) Nevertheless, this study’s participants who died during this 9-month pandemic period were generally elderly and largely chronically ill. Data analysis identified only 4 dead participants who were classified as healthy after their last evaluation. i.e., no relevant regular medications, no relevant clinical diseases diagnosed and normal results on the evaluation protocol – 1 (2%) in non-COVID-19 vs 3 (8%) in COVID-19 groups [p=.518] -.

Coronary artery disease, arterial hypertension and regular use of ß-blockers were present in 52 to 54 (54-56%) of the 96 participants with non-COVID and COVID-19 deaths; more than half of the participants with coronary artery disease also had a previous diagnosis of myocardial infarction. Additionally, dyslipidemia (54%) and diabetes mellitus (32%) were also very common among the participants. On the other hand, it should be noted that the prevalence of obesity, as defined by a body mass index > 30 kg.m^-2^, was relatively low. According to body mass index, 25% of all participants had normal weight/height relationship and only 7 (20%) of those that died from COVID-19 were classified as obese. Moreover, only one participant dying from atherosclerotic heart disease had a body mass index indicating morbid obesity. There are recent data from the UK Biobank(51) suggesting that among those older than 70 years, body mass index tended to be a weaker predictor of the COVID-19 clinical course. Notwithstanding, considering the relatively small sample size of our study, affirmative conclusions could not be made from regarding this matter.

It was not unexpected that resting lung spirometry and cardiac vagal tone were below the age- and sex-predicted reference values, since most participants had chronic non-communicable diseases. It is interesting to note further that despite the fact that most of the participants had a diagnosis of arterial hypertension, the average value for seated pre-CPX and maximal systolic and diastolic blood pressure were in an acceptable range, suggesting that these participants were, in fact, medically well-controlled in terms of arterial hypertension. Most likely due to the fact that the regular use of ß-blockers was highly prevalent among the participants, the mean overall maximal heart rate was only about 78% of the age-predicted value,(53) though it did not differ between the two groups.

Aerobic (cardiorespiratory) fitness variables were objectively measured in 94% of all participants by expired gas analysis during a maximal cycling CPX using an individualized ramp protocol, with an optimal duration having been obtained in the majority of the participants. No statistical differences in cardiorespiratory fitness were observed between groups. The mean overall aerobic fitness as reflected by VO2 max was much lower than age- and sex-predicted values, clearly indicating that the vast majority of participants were aerobically unfit. The cardiorespiratory optimal point was higher than age- and sex-predicted for the majority of the participants and, as defined in a previous epidemiological study, 11%, 53% and 36% of the participants were, respectively, classified as good, average and poor for the cardiorespiratory optimal point when considering previously defined cut-off values for all-cause mortality. While we were unable to find in our literature search papers directly relating aerobic fitness to COVID-19 mortality, some but not all available studies support the idea that high levels of aerobic fitness or physical activity could prevent hospitalizations due to COVID-19.(18, 47, 54, 55). Therefore, some uncertainty remains as recently stated by Zbinden-Foncea et al.(17): “whether a high level of cardiorespiratory fitness can reduce the early amplified proinflammatory responses in patients infected with SARS-CoV-2 and confer some protective effect against the development and severity of the disease remains to be established from retrospective epidemiological data”.

MUSK fitness variables were assessed by four specific tests that were applied according to well-defined protocols by properly trained personnel. Similar to the aerobic-derived CPX variables, no statistical differences were found between non-COVID-19 and COVID-19 death groups. Nevertheless, it should be noted that by using age and sex-reference values for the four tests, it was clear that MUSK fitness was lower than expected in all four tests, expressed in both absolute and relative to body weight (when applicable) for the vast majority of the participants in the non-COVID-19 and COVID-19 death groups.

Considering our previous studies, it is worth adding a comment regarding two other CPX variables – exercise heart rate gradient (56) and maximal oxygen pulse,(57, 58) both previously shown to be associated with all-cause mortality and in principle could have been predictors of mortality in the current study. However, a more recent study has shown that the exercise heart rate gradient was not useful to predict mortality in patients using ß-blockers (59), as was the case for 56% of our participants, and its use was discarded for this study. Regarding maximal absolute or relative oxygen pulse, since VO2 max and maximal heart rate were closely similar in both groups, it was thought that the inclusion of oxygen pulse as a study variable would not be meaningful.

Looking at the individual profile of each participant that has died could be very informative (readers may contact the corresponding author for access to those detailed data). For example, the youngest participant that died was one of the very few that had an undetermined cause of death (R99 code in ICD-10) occurring at 51 years of age. Interestingly, in his last evaluation (the fourth in CLINIMEX) carried out ten years before the death, he presented with arterial hypertension and dyslipidemia and had the best non-age adjusted aerobic indicators - highest VO2 max (37.9 mL.kg-1.min-1) and lowest cardiorespiratory optimal point (19.1) -among all 96 participants that died.

In summary, participants of the CLINIMEX cohort that died during this 9-month pandemic period were primarily elderly and largely chronically ill and unfit. In retrospect, our data suggest that the occurrence of death of any cause seems to be a relatively rare event in a given 9-month period among our healthy middle-aged and elderly cohort participants. In a future study, it would be likely relevant to analyze the aerobic and MUSK fitness levels among elderly CLINIMEX cohort survivors from the pandemic of COVID-19.

### Limitations

This study has several relevant limitations: 1) The study has a retrospective design and there was a large range in the interval of time between the evaluations and the outcomes, i.e., death, as previously mentioned. While recent clinical data was available for some unhealthy and healthy participants that died, as in those that were attending onsite or remote exercise programs shortly before death, regarding many others, no recent information was available for the current study. However, this situation is similar for both COVID-19 and non-COVID-19 deaths in terms of the number of days elapsed between last evaluation and fatal outcomes (see table 2); 2) For this study, we relied on the information made available on the death certificates and no formal attempt was made to check their accuracy; 3) while this paper is being written (mid-February), the pandemic continues strong and Brazil is reaching new tragic records of 7-day rolling average of COVID-19 deaths/day; thus, we do not know if the findings herein would differ if a longer period of observation is considered; 4) A theoretical and logical possibility was that the participants dying from COVID-19 were more exposed and less “prudent” in regard to the virus infection. Unfortunately, there is no available information about how adherent the participants were to recommended sanitary precautions; however, as far as we could find out from information obtained from spouses, relatives and friends of the participants, the vast majority of them adopted a high level of self-protection and this did not seem to vary among participants in the two groups;. 5) While we have been aware of several cases of COVID-19 infection among participants in our cohort, unfortunately, we were unable to obtain reliable data regarding the actual number or percentage of them that got infected or were hospitalized or needed mechanical ventilation due to COVID-19. As such, we were unable to analyze the association between our selected variables and these rather minor or intermediate but clinically relevant health outcomes in our cohort as other studies have done.(18, 19) 6) In relation to the analysis of conventional cardiovascular risk factors, it is worthwhile mentioning the reason why smoking was not considered in the study. In contrast to several other countries in the world, the prevalence of smoking status is currently very low in the Brazilian adult population, and in particular among the elderly CLINIMEX cohort. At baseline, only 1 to 2% of all participants were current smokers, although the majority of them reported to have smoked in the past. Therefore, smoking habit was not considered to be relevant for this study.

### Positive aspects

This study also has several positive aspects or features that should be underscored. 1) Only six experienced Exercise & Sport physicians were involved in all evaluations over the 26 years of data collection, warranting the same protocol and quality to the data obtained for all testing and measurements; 2) All maximal leg cycling exercise tests were conducted in a single exercise lab following individualized ramp protocols and expired gas was collected and analyzed in metabolic units from the same manufacturer; None of our physicians were involved in the care of COVID-19 for the participants in the cohort or in signing their death certificates, which were independently and officially obtained from the State Secretary of Health; 4) The use of data originating from participants from the same cohort for comparisons – non-COVID-19 versus COVID-19 deaths – provided a unique opportunity to address these issues, particularly given the fact that both groups were similar in terms of sex distribution, age at evaluation and at death and percentage of those regularly using ß-blockers.

## Conclusions

COVID-19 was the most frequent and premature cause of death in a convenience sample of primarily white and middle-aged and elderly participants with chronic non-communicable diseases in the CLINIMEX cohort (located in Copacabana – Rio de Janeiro – Brazil). Our data suggest that exercise/sport history and aerobic and MUSK physical fitness testing data obtained some years earlier were unable to distinguish between non-COVID-19 and COVID-19 deaths. However, CLINIMEX cohort participants who died due to non-COVID-19 related causes during the first 9-month period (mid-March to mid-December/2020) of the COVID-19 pandemic were more severely affected by non-communicable diseases or cardiovascular risk factors than those who became victims of COVID-19. Therefore, the presence of cardiovascular risk factors like obesity was associated with an increased number of deaths in the non-COVID-19 but not COVID-19 groups.

### WHAT IS ALREADY KNOWN

- COVID-19 has caused thousands of deaths worldwide and Brazil has been one of the most affected countries.
- COVID-19 deaths are unevenly distributed within countries, regions and cities. As of mid-February/2021, the Copacabana area in the city of Rio de Janeiro has been heavily hit by COVID-19, with a tragic figure of 1 death per 266 inhabitants.
- CLINIMEX, a privately-owned Exercise Medicine Clinic situated in Copacabana, started an open cohort in 1994 that currently has over 9,000 participants and a large amount of accumulated data – including history of exercise and aerobic and musculoskeletal fitness; this cohort has undergone periodic updates of their vital status and mortality information.

### WHAT THIS STUDY ADDS

- COVID-19 is the most frequent cause of death in a population of primarily white, elderly and unfit men and women with various non-communicable diseases
- In comparing clinical, exercise, aerobic and musculoskeletal fitness data obtained from middle-aged and elderly participants from a Rio de Janeiro cohort that died in the first 9-month period of COVID-19 pandemic, those who died from COVID-19 tended to be somewhat healthier than those dying from other causes. However, no differences were identified in sex distribution, history of exercise/sports and several indicators of aerobic and musculoskeletal fitness

### FUTURE RESEARCH NEEDS

- To assess whether or not objectively measured aerobic and musculoskeletal physical fitness is associated with COVID-19 deaths in both healthy and unhealthy young and middle-aged men and women adults
- To verify whether recent levels of physical activity and/or exercise and/or sport are able to predict the clinical course of COVID-19 once infection occurs

## Supporting information

CLINIMEX COHORT COVID-19 DEATH STUDY: STATISTICS FOR ALL 49 VARIABLES

CLINIMEX EXERCISE COHORT_BRIEF DESCRIPTION

CLINIMEX EXERCISE COHORT_EVALUATION PROTOCOL DESCRIPTIONprotocol description_final

## Data Availability

interested readers may request access to more detailed data information directly to contacting author

## Acknowledgements

The authors thank to the Rio de Janeiro State Secretary of Health by allowing us to access to the complete and updated information about vital data

## Author Declarations

This study was authorized by CONEP – Conselho Nacional de Ética em Pesquisa - Brazilian Government and ethics committee of the IPCFEx/Brazilian Army (reference number 4.459.555)

## References

1. Worldometer. https://www.worldometers.info/coronavirus/ 2021 [Available from: https://www.worldometers.info/coronavirus/.

2. Woolf SH, Chapman DA, Lee JH. COVID-19 as the Leading Cause of Death in the United States. JAMA. 2021;325(2):123–4.

3. Pizzo PA, Spiegel D, Mello MM. When Physicians Engage in Practices That Threaten the Nation’s Health. JAMA. 2021. 10.1001/jama.2021.0122.

4. Koh HK, Geller AC, VanderWeele TJ. Deaths From COVID-19. JAMA. 2021;325(2):133-4.

5. Cardoso CRB, Fernandes APM, Santos I. What happens in Brazil? A pandemic of misinformation that culminates in an endless disease burden. Rev Soc Bras Med Trop. 2020;54:e07132020.

6. Abbasi K. Covid-19: Social murder, they wrote-elected, unaccountable, and unrepentant. BMJ. 2021;372:314.

7. Else H. How a torrent of COVID science changed research publishing - in seven charts. Nature. 2020;588(7839):553.

8. Araujo CG. Physical Activity, Exercise and Sports and Covid-19: What Really Matters. Int J Cardiovasc Sci. 2021;34(2):113–5.

9. Pitanga FJG, Beck CC, Pitanga CPS. Should Physical Activity Be Considered Essential During the COVID-19 Pandemic? Int J Cardiovascular Sci. 2020;33(4):401–3.

10. McKinney J, Connelly KA, Dorian P, Fournier A, Goodman JM, Grubic N, et al. COVID-19-Myocarditis and Return to Play: Reflections and Recommendations From a Canadian Working Group. Can J Cardiol. 2020. 10.1016/j.cjca.2020.11.007

11. Salman D, Vishnubala D, Le Feuvre P, Beaney T, Korgaonkar J, Majeed A, et al. Returning to physical activity after covid-19. BMJ. 2021;372:m4721.

12. Faghy MA, Arena R, Stoner L, Haraf RH, Josephson R, Hills AP, et al. The need for exercise sciences and an integrated response to COVID-19: A position statement from the international HL-PIVOT network. Prog Cardiovasc Dis. 2021. 10.1016/j.pcad.2021.01.004.

13. Bhatia RT, Marwaha S, Malhotra A, Iqbal Z, Hughes C, Borjesson M, et al. Exercise in the Severe Acute Respiratory Syndrome Coronavirus-2 (SARS-CoV-2) era: A Question and Answer session with the experts Endorsed by the section of Sports Cardiology & Exercise of the European Association of Preventive Cardiology (EAPC). Eur J Prev Cardiol. 2020;27(12):1242–51.

14. Laukkanen JA, Zaccardi F, Khan H, Kurl S, Jae SY, Rauramaa R. Long-term Change in Cardiorespiratory Fitness and All-Cause Mortality: A Population-Based Follow-up Study. Mayo Clin Proc. 2016;91(9):1183–8.

15. Myers J, Prakash M, Froelicher V, Do D, Partington S, Atwood JE. Exercise capacity and mortality among men referred for exercise testing. N Engl J Med. 2002;346(11):793–801.

16. Brito LB, Ricardo DR, Araujo DS, Ramos PS, Myers J, Araujo CG. Ability to sit and rise from the floor as a predictor of all-cause mortality. Eur J Prev Cardiol. 2014;21(7):892–8.

17. Zbinden-Foncea H, Francaux M, Deldicque L, Hawley JA. Does High Cardiorespiratory Fitness Confer Some Protection Against Proinflammatory Responses After Infection by SARS-CoV-2? Obesity (Silver Spring). 2020;28(8):1378–81.

18. Brawner CA, Ehrman JK, Bole S, Kerrigan DJ, Parikh SS, Lewis BK, et al. Inverse Relationship of Maximal Exercise Capacity to Hospitalization Secondary to Coronavirus Disease 2019. Mayo Clin Proc. 2021;96(1):32–9.

19. Cheval B, Sieber S, Maltagliati S, Millet GP, Formánek T, Chalabaev A, et al. Muscle strength is associated with COVID-19 hospitalization in adults 50 years of age and older. medRxiv. 2021:2021.02.02.21250909.

20. Silva FB, Fonseca B, Domecg F, Facio MR, Prado C, Toledo L, et al. Athletes Health during Pandemic Times: Hospitalization Rates and Variables Related to COVID-19 Prevalence among Endurance Athletes. Int J Cardiovasc Sci. 2021. 10.36660/ijcs.20200208.

21. Salman D, Beaney T, Robb CE, de Jager Loots CA, Giannakopoulou P, Udeh-Momoh C, et al. The impact of social restrictions during the COVID-19 pandemic on the physical activity levels of older adults: a baseline analysis of the CHARIOT COVID-19 Rapid Response prospective cohort study. medRxiv. 2021:2021.01.26.21250520.

22. Mudenda S, Mukosha M, Mwila C, Saleem Z, Kalungia AC, Munkombwe D, et al. Impact of the Coronavirus Disease (COVID-19) on the Mental Health and Physical Activity of Pharmacy Students at the University of Zambia: A Cross-Sectional Study. medRxiv. 2021:2021.01.11.21249547.

23. Fearnbach SN, Flanagan EW, Hochsmann C, Beyl RA, Altazan AD, Martin CK, et al. Factors Protecting against a Decline in Physical Activity during the COVID-19 Pandemic. Med Sci Sports Exerc. 2021. 10.1249/MSS.0000000000002602.

24. Eshelby V, Sogut M, Jolly K, Vlaev I, Elliott MT. Stay Home and Stay Active? The impact of stay-at-home restrictions on physical activity routines in the UK during the COVID-19 pandemic. medRxiv. 2021:2021.01.31.21250863.

25. do Amaral VT, Marçal IR, da Cruz Silva T, Souza FB, Munhoz YV, Camprigher Witzler PH, et al. Home confinement during COVID-19 pandemic reduced physical activity but not health-related quality of life in previously active older women. medRxiv. 2020:2020.12.21.20248662.

26. Stovkwell S, Trott M, Tully M, Shin J, Barnett Y, Butler L, et al. Changes in physical activity and sedentary behaviours from before to during the COVID-19 pandemic lockdown: a systematic review. BMJ Open Sport & Exercise Medicine. 2021;7:e000960.

27. Pecci C, Ajmal M. Cardiac Rehab in the COVID-19 Pandemic. Am J Med. 2021. 10.1016/j.amjmed.2021.01.007.

28. Araujo CG, De Souza e Silva Cg. A multiprofessional face-to-face and remote real-time hybrid exercise-based cardiac rehabilitation: an innovative proposal during COVID-19 pandemic. Can J Cardiol. 2021. 10.1016/j.cjca.2020.12.026.

29. Ghisi GLdM, Xu Z, Liu X, Mola A, Gallagher R, Babu AS, et al. Impacts of the COVID-19 Pandemic on Cardiac Rehabilitation Delivery around the World. medRxiv. 2020:2020.11.11.20230045.

30. Frota AX, Vieira MC, Soares CCS, Silva PSD, Silva G, Mendes F, et al. Functional capacity and rehabilitation strategies in Covid-19 patients: current knowledge and challenges. Rev Soc Bras Med Trop. 2021;54:e07892020.

31. Cavalcante JR, Abreu AJL. COVID-19 in the city of Rio de Janeiro: spatial analysis of first confirmed cases and deaths. Epidemiol Serv Saude. 2020;29(3):e2020204.

32. IBGE. https://www.ibge.gov.br/cidades-e-estados/rj/rio-de-janeiro.html 2021 [Available from: https://www.ibge.gov.br/cidades-e-estados/rj/rio-de-janeiro.html.]

33. Araujo CG, Scharhag J. Athlete: a working definition for medical and health sciences research. Scand J Med Sci Sports. 2016;26(1):4–7.

34. Araújo Cgs. Flexitest: an innovative flexibility assessment method. Champaign: Human Kinetics; 2003.

35. Herdy AH, Ritt LE, Stein R, Araújo CG, Milani M, Meneghelo RS, et al. Cardiopulmonary Exercise Test: Background, Applicability and Interpretation. Arq Bras Cardiol. 2016;107(5):467–81.

36. Balassiano DH, Araujo CG. Maximal heart rate: influence of sport practice during childhood and adolescence. Arq Bras Cardiol. 2013;100(4):333–8.

37. Araujo CG, Nobrega AC, Castro CL. Heart rate responses to deep breathing and 4-seconds of exercise before and after pharmacological blockade with atropine and propranolol. Clin Auton Res. 1992;2(1):35–40.

38. Araujo CG, de Castro Clb, Franca JF, Ramos PS. 4-Second Exercise Test: Reference Values for Ages 18-81 Years. Arquivos Brasileiros De Cardiologia. 2015;104(5):366–73.

39. Vianna LC, Oliveira RB, Araujo CG. Age-related decline in handgrip strength differs according to gender. J Strength Cond Res. 2007;21(4):1310–4.

40. Simão Junior RF, Monteiro WD, Araújo Cgs. Potência muscular máxima na flexão do cotovelo uni e bilateral. Rev Bras Med Esporte. 2001;7:157–62.

41. Coelho CW, Hamar D, de Araujo CG. Physiological responses using 2 high-speed resistance training protocols. J Strength Cond Res. 2003;17(2):334–7.

42. Araujo CG. Flexibility assessment: normative values for flexitest from 5 to 91 years of age. Arq Bras Cardiol. 2008;90(4):257–63.

43. Araújo Cgs, Castro CLB, Franca JFC, Araújo DS. Sitting-rising test: Sex-and age-reference scores derived from 6141 adults. Eur J Prev Cardiol. 2020;27:888–90.

44. Ramos PS, Araújo CG. Cardiorespiratory optimal point during exercise testing as a predictor of all-cause mortality. Rev Port Cardiol. 2017;36(4):261–9.

45. Ramos PS, Ricardo DR, Araujo CG. Cardiorespiratory optimal point: a submaximal variable of the cardiopulmonary exercise testing. Arq Bras Cardiol. 2012;99(5):988–96.

46. Barbosa IR, Galvão MHR, de Souza TA, Gomes SM, Medeiros AA, de Lima KC. Incidence of and mortality from COVID-19 in the older Brazilian population and its relationship with contextual indicators: an ecological study. Rev Bras Ger Gerontol. 2020;23(1):e200171.

47. Ho FK, Celis-Morales CA, Gray SR, Katikireddi SV, Niedzwiedz CL, Hastie C, et al. Modifiable and non-modifiable risk factors for COVID-19: results from UK Biobank. medRxiv. 2020:2020.04.28.20083295.

48. Juul FE, Jodal HC, Barua I, Refsum E, Olsvik Ø, Helsingen LM, et al. Mortality in Norway and Sweden before and after the Covid-19 outbreak: a cohort study. medRxiv. 2020:2020.11.11.20229708.

49. khedr EM, Daef E, Mohamed-Hussein A, Mostafa EF, zein M, Hassany SM, et al. Impact of comorbidities on COVID-19 outcome. medRxiv. 2020:2020.11.28.20240267.

50. O’Gallagher K, Shek A, Bean DM, Bendayan R, Teo JTH, Dobson RJB, et al. Pre-existing cardiovascular disease rather than cardiovascular risk factors drives mortality in COVID-19. medRxiv. 2020:2020.12.02.20242933.

51. Sattar N, Ho FK, Gill JM, Ghouri N, Gray SR, Celis-Morales CA, et al. BMI and future risk for COVID-19 infection and death across sex, age and ethnicity: preliminary findings from UK biobank. medRxiv. 2020:2020.06.05.20122226.

52. Clouston SAP, Luft BJ, Sun E. History of premorbid depression is a risk factor for COVID-related mortality: Analysis of 1,387 COVID+ patients. medRxiv. 2021:2020.12.17.20248362.

53. Duarte CV, Araujo CG. Cardiac vagal index does not explain age-independent maximal heart rate. Int J Sports Med. 2013;34(6):502–6.

54. de Souza FR, Motta-Santos D, Santos Soares Dd, de Lima JB, Cardozo GG, Pinto Guimarães LS, et al. Physical Activity Decreases the Prevalence of COVID-19-associated Hospitalization: Brazil EXTRA Study. medRxiv. 2020:2020.10.14.20212704.

55. Pinto AJ, Goessler KF, Fernandes AL, Murai IH, Sales LP, Reis BZ, et al. No associations between physical activity and clinical outcomes among hospitalized patients with severe COVID-19. medRxiv. 2020:2020.11.25.20237925.

56. Duarte CV, Myers J, de Araujo CG. Exercise heart rate gradient: a novel index to predict all-cause mortality. Eur J Prev Cardiol. 2015;22(5):629–35.

57. Oliveira RB, Myers J, Araujo CG, Abella J, Mandic S, Froelicher V. Maximal exercise oxygen pulse as a predictor of mortality among male veterans referred for exercise testing. Eur J Cardiovasc Prev Rehabil. 2009;16(3):358–64.

58. Laukkanen JA, Araujo CGS, Kurl S, Khan H, Jae SY, Guazzi M, et al. Relative peak exercise oxygen pulse is related to sudden cardiac death, cardiovascular and all-cause mortality in middle-aged men. Eur J Prev Cardiol. 2018;25(7):772–82.

59. Wang S, Muller J, Goeder D, Araujo CG, Silva C, Myers J. Effect of Beta-Blocker Use on Exercise Heart Rate Gradient and Reclassification of Mortality Risk in Patients Referred for Exercise Testing. Am J Cardiol. 2020;130:152–6.

