## Supplementary material for "Mortality during the COVID-19 pandemic: findings from the CLINIMEX exercise cohort in the year of 2020": CLINIMEX EXERCISE COHORT_BRIEF DESCRIPTION

SUPPLEMENTAL MATERIAL

SUPPLEMENTAL METHODS

*CLINIMEX (Exercise Medicine Clinic) Exercise cohort*

Since its opening in January 1994, all individuals that were attended at CLINIMEX signed an informed consent specifically allowing that their data, preserved anonymity, could be used for research studies. Evaluation and medically-supervised exercise program (MSEP) protocols, as well as the research proposal, have been repeatedly evaluated and granted approval by different institutional Ethics in Research committees and is also registered at Plataforma Brasil – CONEP.

The CLINIMEX Exercise cohort comprises all the individuals that have been evaluated in our Clinic since its opening in 1994, and, therefore, it is an open cohort. The information is digitally recorded and available for research. It is worthwhile to underscore that all evaluations over these 27 years were conducted by a small team of Exercise & Sports physicians that strictly adhered a rigid protocol.

The illustration below briefly summarizes the cohort status as it was on December 2020.

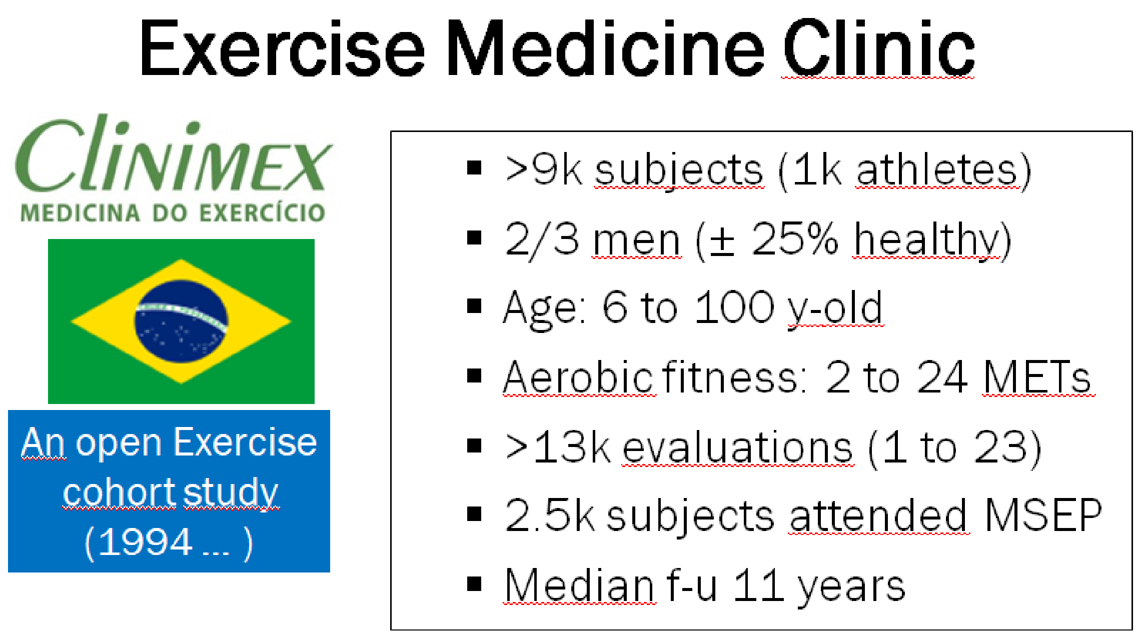
