## Supplementary material for "Mortality during the COVID-19 pandemic: findings from the CLINIMEX exercise cohort in the year of 2020": CLINIMEX EXERCISE COHORT_EVALUATION PROTOCOL DESCRIPTIONprotocol description_final

SUPPLEMENTAL MATERIAL

SUPPLEMENTAL METHODS

### *CLINIMEX (Exercise Medicine Clinic) Evaluation Protocol*

The CLINIMEX evaluation protocol consists in several clinical measurements following a strict protocol developed in our Clinic in order to assess aerobic and non-aerobic physical fitness and to advise/prescribe physical exercise.

#### Clinical history and physical examination

The CLINIMEX evaluation protocol begins with a detailed anamnesis, which includes the assessment of the individual's patterns of physical activity and exercise/sport in different life spans as follows: 1- childhood and adolescence; 2- adult life; and 3- the last 12 months from the day of the evaluation; and previous participation in competitive sport and in supervised exercise programs. For each one of these life spans, an incremental ordinal score from 0 to 4 is attributed, where 0 refers to sedentary lifestyle, 1 represents insufficiently active [less than minimum recommended dose for age], 2 means achieving the age-recommended dose in terms of regular physical activity, exercise and/or sports, 3 reflects a very active lifestyle [exceeding the minimal recommended dose for age], and 4 is attributed for those that are exceeding the minimal recommended dose in several times and/or are involved in competitive sports that have a high aerobic requirement. Next, a physical examination with an emphasis on the cardiovascular system is performed. Also, information on medications of regular use and results of previous laboratory and complementary exams, when available, are digitally recorded.

#### Resting electrocardiogram

Under climatically controlled conditions of temperature (21$^{o}$C to 24$^{o}$C) and humidity (40% to 60%), a standard 12-lead resting electrocardiogram in the supine position is registered. During its registration, respiratory sinus arrhythmia is assessed through the measurement of heart rate variation following a slow maximal inspiration and expiration. The ECG recording is obtained and stored using a digital electrocardiograph and specific Wincardio software (Micromed, Brazil).

Kinanthropometric measurements

Several kinanthropometric measurements are obtained during the CLINIMEX evaluation protocol.

First, basic anthropometry is assessed. Body weight is measured with a Welmy scale, with 0.1 kg resolution; height is measured with a Sanny stadiometer with 0.1 cm resolution; and waist circumference is measured in the upright position, at the umbilicus level, with a Seca or Gullik anthropometric tape with 0.1 cm resolution. Body mass index ($kg/m^2$) and waist-height ratio are calculated.

In sequence, skinfolds are measured in six specific sites (medial calf, anterior thigh, triceptal, subscapular, suprailiac and abdominal) using a skinfold caliper (Skyndex or Harpender, if skinfold >50 mm) at 0.1 mm resolution, bone diameters of humerus and femur are measured by an adapted Mitutoyo caliper with Vernier scale at 0.01 mm, and upper-arm (biceps muscle relaxed and contracted) and calf girths are obtained for calculation of body composition and Heath-Carter anthropometric somatotype.

Next, muscle strength and power are assessed through handgrip strength (absolute and relative to body weight) that is measured twice in both hands with arm completely parallel to the longitudinal axis of the body – the largest result being chosen for subsequent analysis - and maximum power (absolute and relative to body weight) in a standing upright row movement exercise quantified in watts (FitroDyne, Slovakia).

Flexibility is evaluated by applying the Flexitest. Flexitest evaluates the maximal passive range of motion of 20 joint movements. The movement is always initiated at a baseline position towards greater joint amplitude. All the movements are performed passively up to either physical limitation or the individual's complaint of discomfort. For scoring, the range of motion passively obtained for each movement is compared with a reference chart (visual comparison). Discontinuous values ranging from 0 to 4 are assigned to each movement, and no intermediate values are allowed. It is then possible to study the flexibility level in any of 20 individual movements or seven joints. In addition, since flexibility scores for all individual movements present a normal distribution – median of 2 –, it is possible to add them to obtain a global or overall body flexibility dimensionless score, that is called Flexindex (range: 0 to 80). Usually, all movements that are bilateral, by standardization, are performed on the individual's right side.

Balance

Balance is assessed in four different positions in a platform (Fitrosway, Slovakia) as follows: 1) double leg stance with feet together and eyes opened; 2) double leg stance with feet together and eyes closed; 3) single leg stance on right foot; 4) and single leg stance on left foot. The individual must be barefoot and he/she must remain on each position for 10 seconds. The center of pressure displacement in all directions during these 10 seconds for each position is determined and recorded by the software.

Sitting-rising test

The sitting-rising test (SRT) is a safe, costless, quick to apply assessment tool for the evaluation of the four non-aerobic fitness components - flexibility, muscle power, dynamic balance and body composition. The SRT consists in the quantification of the number of supports (hands and/or knees, or hands or forearms on knees) one utilizes in order to sit and to rise from the floor. Independent scores are provided to each of the two actions - sitting and rising. The maximal grade is 5 for each one of the actions, losing one point for each support and additional half point for any detectable unbalance during the performance. A composite score is obtained by adding the sitting and the rising scores, and ranges from 0 to 10 at half point intervals.

Resting lung spirometry

A standard resting lung spirometry – forced vital capacity and flow-volume curve - is also performed to evaluate the individual's main lung volumes and flows (Schiller [Switzerland] or Koko [United States] spirometers). The maneuver that achieves the highest sum of forced vital capacity and forced expiratory volume at the first second from three high-quality maneuvers is chosen to represent the individual's results.

The 4-Second Exercise Test

The aim of the 4-Second Exercise Test (4sET) is to assess the integrity of the cardiac parasympathetic branch of the autonomic nervous system in the initial transient of heart rate (rest-exercise transition) and is based on the physiological vagal withdrawal induced by movement of the limbs. Briefly, the 4sET consists of cycling, as fast as possible, an unloaded cycloergometer, from the fifth to the night second of a maximum inspiratory apnea lasting 12 seconds. The individual being tested should follow four verbal commands at each 4 seconds: a) to get a maximal quick full inspiration, primarily through the mouth; b) to cycle as fast as possible; c) to stop abruptly; and d) to expire normally. During this test, a single electrocardiographic tracing is continuously recorded (usually lead CC5 or CM5) during 35 seconds, at a velocity of 25 mm/s that is initiated 5 seconds before the maximum inspiration command. To determine the magnitude of the cardiac vagal tone, the longest RR interval – the one immediately before, or the first during exercise, whatever is the largest one – and the shortest RR interval during exercise – usually the last one – are identified and measured at 10 ms resolution in the digital tracing. The ratio between these two RR interval durations indicates the cardiac vagal index, a dimensionless variable that is obtained by the 4sET. If a longer RR interval duration is detected after the exhalation, it is also recorded and it reflects a vagal rebound phenomena.

Cardiopulmonary exercise test (CPX)

CPX is conducted in a room that is adequately prepared – personnel, equipments, medications and supplies - to handle with medical emergencies that may arise. While both cycling and treadmill CPX could be performed at CLINIMEX, the vast majority of our CPX are undertaken in a lower limb cycle ergometer Cateye EC-1600 (Cateye, Japan) or Inbrasport CG-04 (Inbrasport, Brazil). When appropriate, mostly in exercisers or athletes used to run, CPX is performed on an Inbrasport ATL-2000 treadmill (Inbrasport, Brazil). Very rarely and for specific reasons, a Monark upper limb cycle ergometer (Monark, Sweden) or an Inbrasport arm ergometer (Inbrasport, Brazil) or a Concept II remoergometer (Concept, United States) are used for CPX. For all CPX, expired gases are collected by the use of a Prevent pneumotacograph (MedGraphics, USA) coupled to a mouthpiece, with concomitant nasal occlusion. The expired gases are measured and analyzed by using a $VO_{2000}$ metabolic analyzer (MedGraphics, USA) daily calibrated for volumes and gas fractions before the first assessment and whenever necessary. Using a mixed chamber concept, expired gases are averaged and read every 10 seconds. For simplicity, these 10-second results are, subsequently, averaged for each minute.

During the CPX, the individual is continuously monitored from resting until five minutes after exercise is completed with a digital electrocardiograph (ErgoPC Elite, versions 3.2.1.5 or 3.3.4.3 or 3.3.6.2, Micromed, Brazil). Heart rate is measured on the eletrocardiographic recording (leads CC5 or CM5) at the end of each minute. Blood pressure is measured by physician’s auscultation every minute on the right arm by using a manual sphygmomanometer during exercise and in the first, second, third and fifth minute of recovery (most frequently on the supine position). After being previously explained to the individual, rate of overall perceived exertion in an ordinal incremental scale from 0 to 10 (half-point allowed) is asked at each minute by the physician in charge to the individual and also recorded.

The CPX is always performed according to an individualized ramp protocol, aimed to achieve voluntary exhaustion at 8 to 12 minute duration. The maximum intensity of the exercise is confirmed by maximum voluntary exhaustion (score 10 in the Borg scale ranging from 0 to 10) represented by the incapacity to continue the effort despite strong verbal encouragement. The characterization of CPX as maximum is also confirmed by the impression of the physician in charge and recorded on the CPX's report. CPX is neither interrupted nor considered maximum based exclusively on the heart rate achieved.

The following variables are also obtained from the CPX: 1- maximum oxygen uptake ($VO_2max$): the highest oxygen uptake value obtained at a given minute of the CPX and expressed as absolute ($L.min^{-1}$) or relative to body weight ($mL.kg^{-1}.min^{-1}$) value or as percent of maximal sex- and age-predicted value – men: 60 – 0.55 x age in years and women: 48 – 0.37 x age in years -; 2- anaerobic threshold (in $mL.kg^{-1}.min^{-1}$ or % of $VO_2max$), determined based on the graphical inspection of data on oxygen consumption ($VO_2$) and ventilation (VE), at which point there is a sudden loss in the linearity of VE curve, whereas $VO_2$ continues to increase linearly with the workload; 3- $O_2$ pulse (absolute or per body weight unit), calculated as $VO_2$/heart rate ratio obtained every 10 seconds; 4- Cardiorespiratory optimal point, a dimensionless variable, obtained by identifying the lowest value of the $VE/VO_2$ ratio measured minute-by-minute during the incremental maximum CPX, regardless of when it occurred; 5- Exercise heart rate gradient (EHRG), an index combining heart rate reserve (maximum heart rate minus heart rate at rest) and heart rate recovery (maximum heart rate minus heart rate one minute post exercise); 6- Oxygen saturation, measured by plethysmography using a finger oximeter and recorded minute-by-minute during exercise and in the first five minutes of post-exercise period of CPX. Additionally, curves kinetics for all variables are also evaluated.
